# Chronic respiratory symptoms and chronic obstructive pulmonary disease following completion of pulmonary tuberculosis treatment in Uganda

**DOI:** 10.1101/2023.09.17.23295686

**Authors:** Martha Namusobya, Felix Bongomin, Mukisa John, Ivan Kimuli, Ahmed Ddungu, Charles Batte, Bruce J. Kirenga

**Affiliations:** Department of Clinical Epidemiology and Biostatistics, School of Medicine, College of Health Sciences, Makerere University, Kampala, Uganda; Department of Medical Microbiology and Immunology, Faculty of Medicine, Gulu University, Gulu, Uganda; Division of Evolution, Infection and Genomics, Faculty of Biology, Medicine and Health, University of Manchester, M13 9PL, Oxford Road, Manchester, United Kingdom; Department of Immunology and Molecular Biology, School of Biomedical Sciences, College of Health Sciences, Makerere University, Kampala, Uganda; Makerere Lung Institute, School of Medicine, College of Health Sciences, Makerere University, Kampala, Uganda; Infectious Diseases Institute, College of Health Sciences, Makerere University, Kampala, Uganda

**Keywords:** Pulmonary tuberculosis, Chronic Respiratory Symptoms, Chronic Obstructive Pulmonary Disease, Spirometry, Uganda

## Abstract

**Background:** Prior pulmonary tuberculosis (PTB) is associated with chronic lung impairment, including chronic obstructive pulmonary disease (COPD). We determined the prevalence and associations of chronic respiratory symptoms and COPD following completion of PTB treatment in Uganda.

**Methods:** Between August 2022 and December 2022, we consecutively enrolled post-PTB patients who had successfully completed PTB treatment between January 2016 and January 2020 at Mulago National Referral Hospital, Kampala, Uganda. Chronic respiratory symptoms were defined as the presence of at least one of the following symptoms lasting for at least three months within a year: cough or sputum production, shortness of breath, chest pain, or wheezing, along with an FEV_1_/FVC < 0.70 on spirometry for the definition of COPD.

**Results:** We enrolled 326 participants (median age 36 years; IQR: 30 — 43), 182 (55.8%) were male, and 123 (37.7%) were living with HIV. Fifty-one (15.6%) participants had chronic respiratory symptoms, 5 (9.8%) having COPD; 4 GOLD I and 1 GOLD II. Chronic respiratory symptoms were 89% lower among participants whose PTB treatment outcome was “completed” (adjusted Odds Ratio (aOR): 0.11, 95% confidence interval (CI): 0.01 – 0.87, p<0.01) and they were 74% lower among those with alcohol use disorders (aOR: 0.26, 95% CI: 0.12 – 0.57, p <0.001). Non-HIV immunosuppressive conditions such as diabetes mellitus and chronic steroid use, were significantly associated with both chronic respiratory symptoms (aOR:7.72, 95% CI 3.13 – 19.04, p<0.001) and COPD (aOR: 8.42, 95% CI: 1.32 — 53.47, p=0.024).

**Conclusion:** Chronic pulmonary symptoms, including COPD, are important and yet under recognized complications of PTB treatment in Uganda. Therefore, screening and management in key sub-groups, such as those with immunosuppressive condition, will improve morbidity and quality of life in this population.

## Introduction

Pulmonary tuberculosis (PTB) is associated with parenchymal destruction leading to alteration in lung architecture and chronic lung impairment, including chronic obstructive pulmonary disease (COPD). Many patients remain symptomatic even after ‘successful’ PTB treatment (1). Studies done in Brazil and Malawi showed that prevalence of chronic respiratory symptoms after PTB cure were 45% (2) and 30.7% (3), respectively. The PLATINO study showed that the overall prevalence of airflow obstruction is over 2-folds higher among subjects with a history of tuberculosis(TB), compared to those without TB history (4). In this study, a history of prior TB was associated with a more severe form of COPD (4).

COPD is one of the common complications of PTB (1,6,8). A cross-sectional study done among cured Ugandan drug-resistant (DR) TB patients found a 23% prevalence of COPD(5). This study didn’t include drug-sensitive (DS) TB patients who comprise the majority of TB patients. Another study in Ethiopia showed the prevalence of COPD among successfully treated PTB patients (both DS and DR) to be 41.4% (6). The symptoms of COPD (chronic cough, dyspnea, frequent respiratory infections, fatigue, and wheezing) are similar to PTB symptoms and may plague TB patients long after cure. This may be misdiagnosed as a TB recurrence, and managed inappropriately. This may in turn negatively impact patients’ quality of life, leading to increased morbidity, hospital visits, and mortality (7).

COPD is the third leading cause of death worldwide (11), causing 3.23 million deaths in 2019 (13). Over 80% of these deaths occurred in low- and middle-income countries (LMICs) (8). COPD causes persistent and progressive respiratory symptoms, including difficulty in breathing, cough, and/or phlegm production (8). A study conducted in Malawi showed that at PTB completion, 60.7% of participants reported respiratory symptoms, and 34.2% had abnormal spirometry(3). With such staggering statistics, and paucity of data in our own setting, Uganda; it will be key to not only quantify the burden of post-TB persistent respiratory symptoms and COPD, but also tease out their associations. COPD is manageable, and therefore early diagnosis in this special population will be key in reducing morbidity, mortality, and informing management to improve long term outcomes (9, 10).

The TB burden in Uganda remains high with a prevalence of 197 per 100,000 population (11), thus predisposing many patients to the risk of developing chronic lung disease. TB sequelae include obstructive and restrictive lung diseases, bronchiectasis, pleural disease among others (5, 12). Therefore, we hypothesized that COPD is highly prevalent among Ugandan adults who have chronic pulmonary symptoms after PTB treatment. The aim of this study was threefold: to determine the prevalence of chronic respiratory symptoms after PTB treatment, assess the prevalence of COPD among participants with these symptoms following successful PTB treatment, and identify factors associated with chronic respiratory symptoms and COPD in this population.

## METHODS

### Study design

We conducted a cross-sectional study at the National TB Control Center at Mulago National Referral Hospital (MNRH), Kampala, Uganda between 1^st^ August 2022 and 30^th^ November 2022.

### Study Setting

The TB Unit at MNRH serves as the national TB treatment center in Uganda. The unit uses a mixed model of care, whereby, 1) very sick patients are hospitalized at the start of their TB treatment until clinically stable, and 2) outpatient care where patients continue treatment from the community under supervision. The unit manages about 1,500 TB patients annually, making it the largest treatment center in the country.

### Study population

We enrolled post-PTB patients, both drug-sensitive (DS) and drug resistant (DR), 18 years and older, who cured or completed treatment between January 2016 and January 2020, and whose PTB treatment enrollment dates fell in the 2 randomly selected quarters of each cohort year. Pregnant women, critically ill people, those previously treated for only extra-pulmonary TB, those with active TB (relapses), and those with history of heart disease were excluded.

### Sample size estimation

Using previous data of a 30.7% prevalence of persistent pulmonary symptoms in the post-PTB population (3), at 95% confidence interval (CI), power of 80% and type I error of 5%, a sample size of 327 participants was estimated.

### Study procedure

For all eligible participants, a trained medical officer collected data on clinical and demographic characteristics such as age, sex, HIV status and other key co-morbidities e.g., asthma, clinical symptoms, alcohol and tobacco use, type of fuel used for cooking, the type of infrastructure in which cooking happens, occupation, income, and education status using a standardized structured questionnaire. Participants were consecutively recruited until required sample size was reached.

### Spirometry

Participants with chronic respiratory symptoms as ascertained from the questionnaire had spirometry done to assess for COPD. Spirometry was performed by a trained and experienced spirometry technician and read and interpreted by a pulmonologist at the Makerere University Lung Institute (MLI). MLI is a center of excellence in innovative lung health research that integrates disease prevention, clinical care and training.

COPD can be estimated with a high diagnostic accuracy using spirometry (13, 14). The sensitivity for diagnosing airway obstruction in COPD using spirometry is 92% and specificity is 84% (15). The positive predictive value (PPV) is 63%; negative predictive value (NPV) is 97% (15). A spirometry value of FEV_1_/FVC < 0.70 confirmed the presence of persistent airflow limitation (COPD) (16). The severity of COPD was classified using the Global Initiative for Chronic Obstructive Lung Disease (GOLD) classification (17).

### Data analysis

Participant characteristics that were in form of continuous data were described using medians and interquartile ranges. Categorical data was presented in frequencies, proportions and percentages. Comparison between the different categories (e.g. chronic respiratory symptoms vs no chronic respiratory symptoms and COPD vs no COPD) were made using chi square test while continuous variable comparisons were done using the Wilcoxon Rank Sum test. Pie charts and box plots were drawn for data visualization.

The prevalence of chronic respiratory symptoms after successful PTB treatment was presented as proportions/percentages of individuals with any of the symptoms as per the operational definition. Subgroup analysis was performed considering gender as a possible cause of difference in prevalence of chronic respiratory symptoms.

The prevalence of COPD among participants with chronic respiratory symptoms after successful PTB treatment was determined by spirometry evaluation. This was categorized as a dichotomous variable and data was presented as percentages and pie charts. Subgroup analysis was performed considering gender as a possible cause of difference in prevalence of COPD.

To determine factors associated with chronic respiratory symptoms after successful PTB treatment, the presence of chronic respiratory symptoms was dichotomized into a 1 “yes” 0 “No” variable. We tested for assumptions for collinearity and outliers before running a logistic regression model. A bivariate analysis was carried out between the study independent variables and the presence of chronic respiratory symptoms. Factors with biological plausibility from literature like age and sex and those with P< 0.25 will be considered for further multivariable regression analysis. Interaction will be assessed by forming two-way interaction terms and using the likelihood ratio test to assess significance of the interaction terms. Confounding will be assessed by considering a change of 10% percent or more in the odds ratios as confounding, when a model with the variable and one without it are evaluated. The goodness of fit of the final model will be assessed using the Hosmer-Lemeshow goodness of fit statistics. The odds ratios and their 95 % confidence intervals will be presented. A p<.05 will be considered statistically significant. STATA version 14.0 was used for data analysis.

To determine factors associated COPD after successful PTB treatment, the presence of COPD was dichotomized into a 1 “yes” 0 “No” variable and the process of logistic regression above was repeated but this time with COPD as the outcome variable.

### Ethics

The Makerere University School of Biomedical Science Research and Ethics Committee (SBS-2022-131) and the Uganda National Council for Science and Technology (HS2232ES) approved the study protocol. All study participants provided informed written consent. The authors confirm that the ethical policies of the journal, as noted on the journal’s author guidelines page, have been adhered to.

## Results

### Baseline characteristics of the participants

Of the 326 participants enrolled, 182 (55.8 %) were male with a median age for all participants of 36 (IQR: 30 — 43) years. One hundred and twenty-three (37.7%) participants were living with HIV. Majority of the participants, 206 (63.2%), had received Category 1 PTB treatment. A significant number of participants, 145 (44.5%), had history of alcohol use and 66 (20.2%) had a history of smoking tobacco. Twenty-six (8%) of participants had history of other immunosuppressive conditions other than HIV (particularly diabetes and chronic steroid use).

Thirteen (4%) of participants reported history of childhood measles. There were 6 (1.8%) participants with history of asthma. Four (1.2%) participants had history of severe childhood respiratory infections e.g., pneumonia. Only 1 (0.3%) participant had history of COPD, lung cancer and sarcoidosis each. Majority of participants, 274 (84%), used charcoal as their main source of cooking fuel, whilst majority (46.3%) cooked in kitchens attached to the main house. (**Table 1**).

**Table 1.** Baseline characteristics.

| Characteristic | Frequency | Percentage % | Median and IQR |
| --- | --- | --- | --- |
| **Sex** |  |  |  |
| male | 182 | 55.8 |  |
| female | 144 | 44.2 |  |
| **Age** |  |  |  |
| Median, IQR |  |  | 36 (30-43) |
| <40 | 215 | 65.9 |  |
| 40-49 | 78 | 23.9 |  |
| 50 and above years | 33 | 10.2 |  |
| **PTB treatment regimen/ category** |  |  |  |
| Category 1 | 206 | 63.2 |  |
| Category 2 | 1 | 0.3 |  |
| Category 4 (DR TB treatment) | 119 | 36.5 |  |
| **HIV diagnosis** |  |  |  |
| Positive | 123 | 37.7 |  |
| Negative | 200 | 61.3 |  |
| Unknown | 3 | 0.9 |  |
| **Mycobacteria other than tuberculosis history** |  |  |  |
| Yes | 1 | 0.3 |  |
| No | 325 | 99.7 |  |
| **Asthma history** |  |  |  |
| Yes | 6 | 1.8 |  |
| No | 320 | 98.2 |  |
| **COPD history** |  |  |  |
| Yes | 1 | 0.3 |  |
| No | 325 | 99.7 |  |
| **Sarcoidosis history** |  |  |  |
| Yes | 1 | 0.3 |  |
| No | 325 | 99.7 |  |
| **Lung cancer history** |  |  |  |
| Yes | 1 | 0.3 |  |
| No | 325 | 99.7 |  |
| **Chicken pox history** |  |  |  |
| Yes | 7 | 2.1 |  |
| No | 319 | 97.9 |  |
| **Alcoholism history** |  |  |  |
| Yes | 145 | 44.5 |  |
| No | 181 | 55.5 |  |
| **Any other immunosuppressive conditions other than HIV (e.g. diabetes mellitus, and chronic steroid use) history** |  |  |  |
| Yes | 26 | 8 |  |
| No | 300 | 92 |  |
| **Smoking history** |  |  |  |
| Yes | 66 | 20.2 |  |
| No | 260 | 79.8 |  |
| **Measles history** |  |  |  |
| Yes | 13 | 4 |  |
| No | 313 | 96 |  |
| **History of severe childhood respiratory infections e.g. pneumonia** |  |  |  |
| Yes | 4 | 1.2 |  |
| No | 322 | 98.8 |  |

### Prevalence of chronic respiratory symptoms

Of 326 participants, 51 (15.6%) reported at least one of the chronic respiratory symptoms as per case definition. Forty-eight (14.7%) reported shortness of breath, 31 (9.5%) reported cough, 30 (9.2%) reported chest pain, while 22 (6.7%) reported wheezing, (**Table 2**).

**Table 2.** Prevalence of chronic respiratory symptoms.

| Characteristic | Total, N(%) | Male N(%) | Female, N(%) | P value |
| --- | --- | --- | --- | --- |
| **Persistent cough** |  |  |  |  |
| Yes | 31 (9.5) | 13 (7.1) | 18 (12.5) |  |
| No | 295 (90.5) | 169 (92.9) | 126 (87.5) | 0.102 |
| **Shortness of breath** |  |  |  |  |
| Yes | 48 (14.7) | 21 (11.5) | 27 (18.8) |  |
| No | 278 (85.3) | 161 (88.5) | 117 (81.3) | 0.068 |
| **Chest pain** |  |  |  |  |
| Yes | 30 (9.2) | 14 (7.7) | 16 (11.1) |  |
| No | 296 (90.8) | 168 (92.3) | 128 (88.9) | 0.289 |
| **Wheezing** |  |  |  |  |
| Yes | 22 (6.7) | 6 (3.3) | 16 (11.1) |  |
| No | 304 (93.3) | 176 (96.7) | 128 (88.9) | 0.005 |
| **No symptoms** | 275 (83.4) | 162 (87.9) | 112 (77.8) | 0.015 |

At multivariable analysis, factors that were statistically significantly associated with chronic respiratory symptoms were: participants whose successful PTB treatment outcome was “completed treatment” (aOR: 0.11, 95%CI: 0.01 — 0.87, p=0.037), having any other immunosuppressive conditions other than HIV (e.g. diabetes mellitus, and chronic steroid use) (aOR: 7.72, 95%CI: 3.13 — 19.04, p<0.001), and a history of alcoholism (aOR: 0.26, 95%CI: 0.12 — 0.57, p=0.001), (**Table 3**).

**Table 3.** Multivariate analysis of the independent variables to determine their association with presence of chronic pulmonary symptoms.

| Characteristic | Adjusted odds ratio and 95 % CI | P value |
| --- | --- | --- |
| **PTB treatment outcome** |  |  |
| Cured | Ref |  |
| Completed | 0.11 (0.01-0.87) | 0.037 |
| **Any other immunosuppressive conditions other than HIV (e.g. Diabetes mellitus)** |  |  |
| No | Ref |  |
| Yes | 7.72(3.13-19.04) | <0.001 |
| **Alcoholism** |  |  |
| No | Ref |  |
| Yes | 0.26 (0.12-0.57) | 0.001 |
| **Age in completed years** |  |  |
| <40 | Ref |  |
| 40-49 | 1.54(0.72-3.33) | 0.268 |
| 50 and above years | 1.90 (0.67-5.37) | 0.226 |

### Prevalence of COPD

In the 51 (15.6%) participants with chronic respiratory symptoms in whom spirometry was indicated, 5 (9.8%) had COPD. Of the 5 participants with COPD, 4 (80%) had a classification of GOLD I while 1 (20%) had a classification of GOLD II. Other ventilatory defects on spirometry were: asthma (n=10, 19.6%), and restrictive lung disease (n=3, 5.9%), **Table 4**.

**Table 4.** Prevalence of COPD and other findings on spirometry. N=51.

| Characteristic | Frequency | Percentage |
| --- | --- | --- |
| COPD | 5 | 9.8 |
| Asthma | 10 | 19.6 |
| Restriction | 3 | 5.9 |
| Normal spirometry | 33 | 64.7 |

At multivariable analysis, COPD was independently associated with having other immunosuppressive conditions other than HIV (e.g., diabetes mellitus, and chronic steroid use) (aOR: 8.42, 95%CI: 1.32 — 53.47, p=0.024), (**Table 5**).

**Table 5.** Multivariate analysis of the independent variables to determine their association with presence of COPD.

| Characteristic | Adjusted odds ratio and 95 % CI | P value |
| --- | --- | --- |
| **Other immunosuppressive conditions other than HIV (e.g. diabetes mellitus, chronic steroid use) history** |  |  |
| No | Ref |  |
| Yes | 8.42 (1.32-53.47) | 0.024 |
| **Where do you cook from?** |  |  |
| Kitchen attached to main house | Ref |  |
| Kitchen separate from the main house | 0.88 (0.09-9.00) | 0.915 |
| Outdoors/other | 0.38 (0.04-3.78) | 0.409 |

## Discussion

In this study, we found the prevalence of chronic respiratory symptoms among successfully treated post-TB patients to be 15.6%. Our findings are close in keeping with literature that has reported post-TB lung sequalae prevalence to range anywhere from 18 – 87% (18) among successfully treated PTB patients.

The prevalence of COPD among those with chronic respiratory symptoms was about 10%. This prevalence is lower than the 23% that was found when a cross-sectional study was conducted among cured Ugandan DR TB patients (5). This may likely be because the study recruited only DR TB patients who make up the biggest population of PTB treatment failures and retreatments (19) and who thus have worse lung sequalae and higher COPD prevalence.

Chronic respiratory symptoms were independently associated with having other immunosuppressive conditions other than HIV (e.g., diabetes mellitus, and chronic steroid use). It’s quite unsurprising that immunosuppressive states would cause chronic respiratory symptoms because PTB is a driver for opportunist fungal pathogens e.g. chronic pulmonary aspergillosis, which flourish in the immunosuppressed host to cause chronic pulmonary symptoms (20, 21).

COPD was also independently associated with immunosuppressive conditions other than HIV e.g., diabetes mellitus and chronic steroid use. Diabetes has been shown to have adverse inflammatory effects on lung anatomy and physiology (22) and whose impact is inadvertently amplified in conditions of concurrent lung damage like post-TB sequalae. Part of COPD management is use of systemic or inhaled steroids which improve symptoms (23) and this might be why we saw an association with chronic steroid use. That possibly to relieve symptoms, these post-TB patients had steroids prescribed whether by self or health workers, even in the absence of a definitive COPD diagnosis, to improve their quality of life.

Despite a relatively high burden of chronic respiratory symptoms among the study participants, COPD only contributed to about 10% of the etiology of these symptoms. Therefore, multiple etiologies may explain chronic respiratory symptoms in the settings of post-PTB treatment. In this study, we were able to show that asthma and restrictive lung disease contributed to an additional 25% of participants with chronic respiratory symptoms. We have previously shown that chronic pulmonary aspergillosis (CPA) occurs in up to 20% of PTB patients with persistent symptoms (24), significantly affecting their quality of life during PTB treatment (25). Therefore, investigations for both infectious complications and ventilatory defects are indicated in patients with chronic respiratory symptoms following completion of PTB treatment.

This study has some limitations. Firstly, this was a single center study, involving mainly patients from the central region of Uganda and may not be representative of other populations since epidemiological patterns may differ across populations. Secondly, we did not perform chest imaging such as chest x-ray or CT scan, which would be helpful to rule out other complications of post-PTB such as CPA and other differential diagnosis. In addition, further evaluation including sputum examination to rule out TB relapse and other intercurrent pulmonary infections were not done. However, we conducted a rigorous respiratory investigation including spirometry, which is the gold standard for COPD diagnosis. Future, prospective, multicenter studies are recommended to substantiate our findings and provide further insides into the pathogenesis of chronic respiratory

## Conclusions

In conclusion, we demonstrated that chronic respiratory symptoms and COPD where of concern in the post-TB patient population, particularly those with history of non-HIV immunosuppressive conditions such as diabetes mellitus and chronic steroid use. COPD is manageable, and therefore early diagnosis in such special populations will be key in reducing morbidity, mortality, and informing management to improve long term outcomes. Additionally, beyond just COPD, other conditions like fungal pathogens may plague post-TB patients and cause chronic respiratory symptoms. These too should be screened for and appropriately managed among this patient population for improved outcomes and quality of life.

## Data Availability

All relevant data are within the manuscript and its Supporting Information files.

## Acknowledgement

We thank the MNRH TB unit for hosting the study. We’re grateful to the study team based there who were key in participant identification and recruitment. We’re grateful to MLI who performed the study spirometry.

